# Molnupiravir, an Oral Antiviral Treatment for COVID-19

**DOI:** 10.1101/2021.06.17.21258639

**Authors:** William Fischer, Joseph J. Eron, Wayne Holman, Myron S. Cohen, Lei Fang, Laura J. Szewczyk, Timothy P Sheahan, Ralph Baric, Katie R. Mollan, Cameron R. Wolfe, Elizabeth R. Duke, Masoud M. Azizad, Katyna Borroto-Esoda, David A. Wohl, Amy James Loftis, Paul Alabanza, Felicia Lipansky, Wendy P. Painter

**Author notes:** Corresponding author: Wendy P. Painter, MD, MPH, Ridgeback Biotherapeutics, Miami, FL, USA.

## Abstract

**Background:** Easily distributed oral antivirals are urgently needed to treat coronavirus disease-2019 (COVID-19), prevent progression to severe illness, and block transmission of severe acute respiratory syndrome coronavirus 2 (SARS-CoV-2). We report the results of a Phase 2a trial evaluating the safety, tolerability, and antiviral efficacy of molnupiravir in the treatment of COVID-19 (ClinicalTrials.gov NCT04405570).

**Methods:** Eligible participants included outpatients with confirmed SARS-CoV-2 infection and symptom onset within 7 days. Participants were randomized 1:1 to 200 mg molnupiravir or placebo, or 3:1 to molnupiravir (400 or 800 mg) or placebo, twice-daily for 5 days. Antiviral activity was assessed as time to undetectable levels of viral RNA by reverse transcriptase polymerase chain reaction and time to elimination of infectious virus isolation from nasopharyngeal swabs.

**Results:** Among 202 treated participants, virus isolation was significantly lower in participants receiving 800 mg molnupiravir (1.9%) versus placebo (16.7%) at Day 3 (p = 0.02). At Day 5, virus was not isolated from any participants receiving 400 or 800 mg molnupiravir, versus 11.1% of those receiving placebo (p = 0.03). Time to viral RNA clearance was decreased and a greater proportion overall achieved clearance in participants administered 800 mg molnupiravir versus placebo (p = 0.01). Molnupiravir was generally well tolerated, with similar numbers of adverse events across all groups.

**Conclusions:** Molnupiravir is the first oral, direct-acting antiviral shown to be highly effective at reducing nasopharyngeal SARS-CoV-2 infectious virus and viral RNA and has a favorable safety and tolerability profile.

## Introduction

Severe acute respiratory syndrome (SARS) coronavirus 2 (SARS-CoV-2), the virus responsible for coronavirus disease-2019 (COVID-19), has caused more than 166,000,000 confirmed infections and 3,400,000 deaths worldwide as of 23 May 2021.^1^

Studies have shown associations between high SARS-CoV-2 nasopharyngeal ribonucleic acid (RNA) levels and both hospitalization rates and infectious (replication competent) virus isolation.^2–7^ Additionally, animal studies found similar associations between viral RNA levels and transmission.^8^ However, no therapies have been shown to eliminate infectious virus and prevent transmission. Thus, there is an urgent need for oral antiviral therapies that can be easily distributed on a scale that meets global demand, reduce disease progression, and prevent SARS-CoV-2 transmission.

Molnupiravir, the prodrug of the ribonucleoside analog β-D-N^4^-hydroxycytidine (NHC), is rapidly converted in plasma to NHC and then to the active 5′-triphosphate form by host kinases.^9^ The active 5′-triphosphate serves as a competitive substrate for virally-encoded RNA-dependent RNA polymerase (RdRp), and once incorporated into nascent viral RNA, induces an antiviral effect via accumulation of mutations that increase with each viral replication cycle.^9–11^ Preclinical studies reveal broad-spectrum antiviral activity against coronaviruses, including SARS-CoV-2 with a high barrier to resistance.^12,13^ In humanized mouse models, molnupiravir treatment and prophylaxis reduced SARS-CoV, SARS-CoV-2, high-risk SARS-like bat coronaviruses, and Middle East respiratory syndrome coronavirus (MERS-CoV) virus replication and pathogenesis.^14,15^ In a SARS-CoV-2 ferret model, molnupiravir treatment completely blocked transmission to untreated animals, emphasizing that early treatment could potentially prevent secondary spread of SARS-CoV-2.^8^

Molnupiravir has been shown to be safe and well tolerated in a first-in-human, Phase 1, trial in healthy volunteers.^9^ We report the results of a Phase 2a, double-blind, placebo-controlled, randomized, multicenter trial designed to evaluate the safety, tolerability, and antiviral activity of molnupiravir dosed twice-daily for 5 days in the treatment of patients with mild to moderate COVID-19. Hypotheses included that molnupiravir would decrease the time to clearance of SARS-CoV-2 virus in nasopharyngeal swabs and be safe and well tolerated in symptomatic SARS-CoV-2-infected adults.

## Methods

### Trial Design and Conduct

Adults aged ≥18 years were eligible if they tested positive for SARS-CoV-2 infection within 96 hours and had symptoms of COVID-19 within 7 days of treatment initiation (Day 1). Antiviral activity, safety, and tolerability were assessed for 28 days following study treatment initiation. Nasopharyngeal swabs were collected on Days 1 (baseline), 3, 5, 7, 14, and 28 for measurement of antiviral activity by reverse transcriptase polymerase chain reaction (RT-PCR) and infectious virus isolation. Safety laboratories were assessed on Days 1, 3, 5, 7, 14, and 28 and adverse events were monitored throughout.

The protocol was approved by WCG institutional review board and written informed consent was obtained from each participant prior to study entry. The study design and analyses are detailed in supplementary information.

### Randomization and Intervention

Participants were randomized 1:1 to 200 mg molnupiravir or matching placebo or 3:1 to molnupiravir (400 or 800 mg) or placebo. Doses were administered orally twice-daily for 5 days and dose escalations occurred following review of safety and virology data from this and other studies of molnupiravir.

### Antiviral Efficacy

The primary antiviral efficacy outcome was time to viral RNA clearance, as measured by quantitative RT-PCR of nasopharyngeal swabs using a laboratory-developed test based on the Centers for Disease Control and Prevention 2019-nCoV emergency use authorization assay. Time to viral RNA clearance was defined as the first timepoint where viral RNA was achieved and was below the limit of quantitation (<1,018 copies/mL). Secondary antiviral efficacy outcomes were time to infectious virus elimination from nasopharyngeal swabs and median viral RNA change from baseline on Days 3, 5, and 7. Infectious virus isolation was performed using Vero C1008 cells and assessed by quantitative RT-PCR of viral RNA in culture medium at 2 and 5 days post inoculation.^3,16^ Samples were considered positive if the supernatant had at least 1,000 copies/mL SARS-CoV-2 RNA at 2 days post inoculation or if the relative fold-change in RNA copy number from cultures days 2 to 5 days post inoculation was >5. Next generation sequencing of RdRp was performed on samples collected on Day 5 to analyze nucleotide changes and confirm the mechanism of action.

### Safety and Tolerability

The primary safety and tolerability objectives assessed adverse events that were Grade 3 or higher and those that led to early treatment discontinuation.

### Clinical and Immunologic Outcomes

Severities and durations of self-reported COVID-19 symptoms (a secondary objective) were collected daily using a symptom diary. Plasma samples were collected to evaluate immune response and serology using antigen-capture enzyme-linked immunosorbent assays.^17–19^

### Statistical Analyses

Time to response for viral RNA negativity was summarized using Kaplan-Meier methodology. Median time to response and cumulative probability of response by visit (with 95% confidence interval) was analyzed by treatment group. Comparisons of treatment effects were performed using log-rank tests. The number and percentage of subjects who were negative for infectious virus isolation were summarized and between-group comparisons were conducted using Fisher’s exact test. Dose-response assessments were performed using the exact Cochran-Armitage trend test.

Treatment comparisons between active drug and placebo groups for SARS-CoV-2 nasopharyngeal viral load change from baseline were analyzed using a mixed model for repeated measures, with restricted maximum likelihood estimation and an unstructured covariance matrix. The model included fixed effects of treatment, study visit, days since COVID-19 symptom onset, and baseline SARS-CoV-2 viral load (log_10_ copies/mL); and interaction terms of treatment by visit, days since COVID-19 symptom onset by visit, and baseline SARS-CoV-2 viral load by visit. The estimated mean treatment difference for active minus placebo at each visit is presented with the 95% confidence interval and corresponding p-value. Comparisons of next-generation sequencing data between treatments were performed using a two-sample t-test, based on the average number of treatment-emergent nucleotide changes. Analyses were conducted using SAS Version 9.4 (SAS Institute Inc., Cary NC) and two-sided tests were performed using an alpha of 0.05 for treatment comparisons. Adjustments for multiple testing were not performed.

## Results

### Participant Demographics and Clinical Characteristics

Two hundred and four participants were randomized and 202 received at least 1 dose of molnupiravir or placebo between 19 June 2020 and 25 January 2021 at 10 sites in the USA (Figure 1; Table 1). Seven participants discontinued participation due to adverse events, lost to follow-up, physician-decision/participant withdrawal. Baseline SARS-CoV-2 antibody prevalence was greater in the 800 mg molnupiravir group (35.3%) compared to the placebo group (18.2%; Table 1).

**Table 1.**
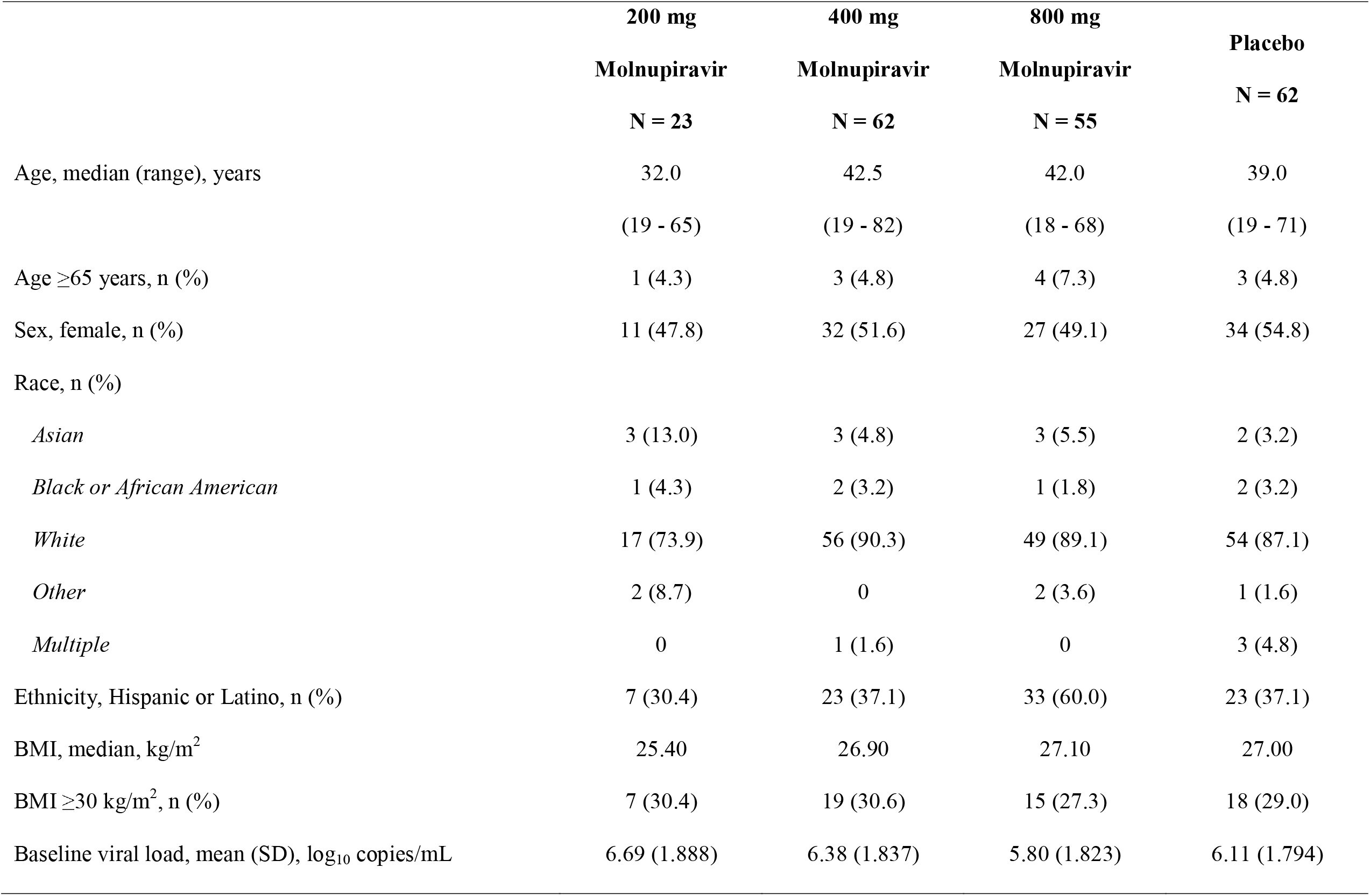
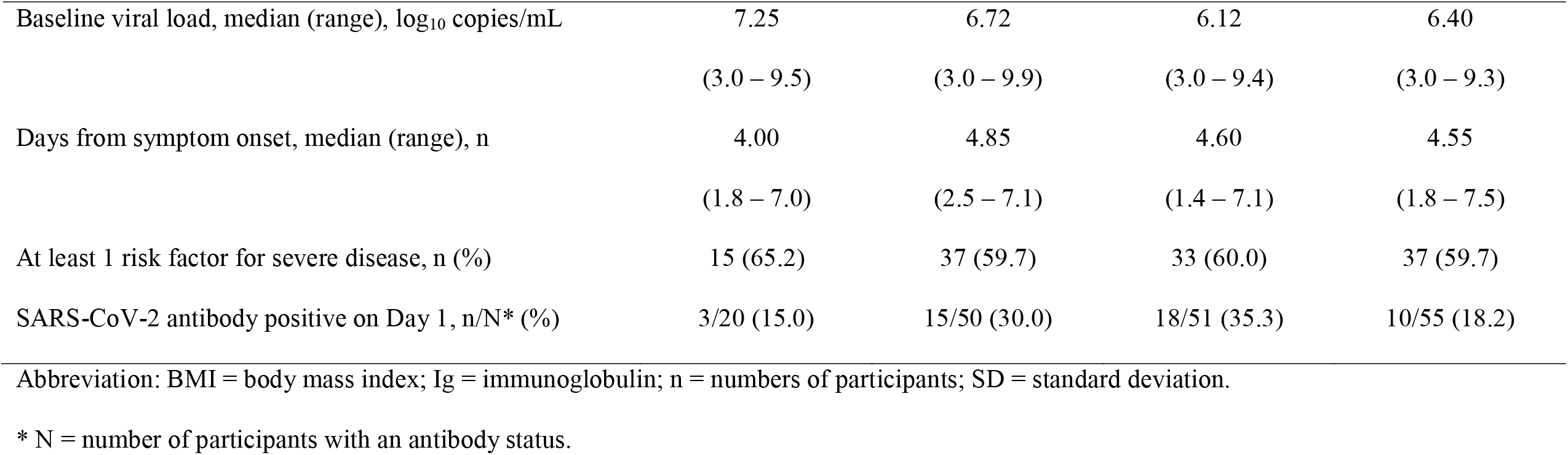
Demographics and Baseline Characteristics.

|  | **200 mg** | **400 mg** | **800 mg** | **Placebo** |
| --- | --- | --- | --- | --- |
|  | **Molnupiravir** | **Molnupiravir** | **Molnupiravir** |  |
|  | **N = 23** | **N = 62** | **N = 55** | **N = 62** |
| Age, median (range), years | 32.0 | 42.5 | 42.0 | 39.0 |
|  | (19 - 65) | (19 - 82) | (18 - 68) | (19 - 71) |
| Age ≥65 years, n (%) | 1 (4.3) | 3 (4.8) | 4 (7.3) | 3 (4.8) |
| Sex, female, n (%) | 11 (47.8) | 32 (51.6) | 27 (49.1) | 34 (54.8) |
| Race, n (%) |  |  |  |  |
| *Asian* | 3 (13.0) | 3 (4.8) | 3 (5.5) | 2 (3.2) |
| *Black or African American* | 1 (4.3) | 2 (3.2) | 1 (1.8) | 2 (3.2) |
| *White* | 17 (73.9) | 56 (90.3) | 49 (89.1) | 54 (87.1) |
| *Other* | 2 (8.7) | 0 | 2 (3.6) | 1 (1.6) |
| *Multiple* | 0 | 1 (1.6) | 0 | 3 (4.8) |
| Ethnicity, Hispanic or Latino, n (%) | 7 (30.4) | 23 (37.1) | 33 (60.0) | 23 (37.1) |
| BMI, median, kg/m<sup>2</sup> | 25.40 | 26.90 | 27.10 | 27.00 |
| BMI ≥30 kg/m<sup>2</sup>, n (%) | 7 (30.4) | 19 (30.6) | 15 (27.3) | 18 (29.0) |
| Baseline viral load, mean (SD), log<sub>10</sub> copies/mL | 6.69 (1.888) | 6.38 (1.837) | 5.80 (1.823) | 6.11 (1.794) |
| Baseline viral load, median (range), log<sub>10</sub> copies/mL | 7.25<br>(3.0 – 9.5) | 6.72<br>(3.0 – 9.9) | 6.12<br>(3.0 – 9.4) | 6.40<br>(3.0 – 9.3) |
| Days from symptom onset, median (range), n | 4.00<br>(1.8 – 7.0) | 4.85<br>(2.5 – 7.1) | 4.60<br>(1.4 – 7.1) | 4.55<br>(1.8 – 7.5) |
| At least 1 risk factor for severe disease, n (%) | 15 (65.2) | 37 (59.7) | 33 (60.0) | 37 (59.7) |
| SARS-CoV-2 antibody positive on Day 1, n/N* (%) | 3/20 (15.0) | 15/50 (30.0) | 18/51 (35.3) | 10/55 (18.2) |

Abbreviation: BMI = body mass index; Ig = immunoglobulin; n = numbers of participants; SD = standard deviation.
\* N = number of participants with an antibody status.

**Figure 1.**
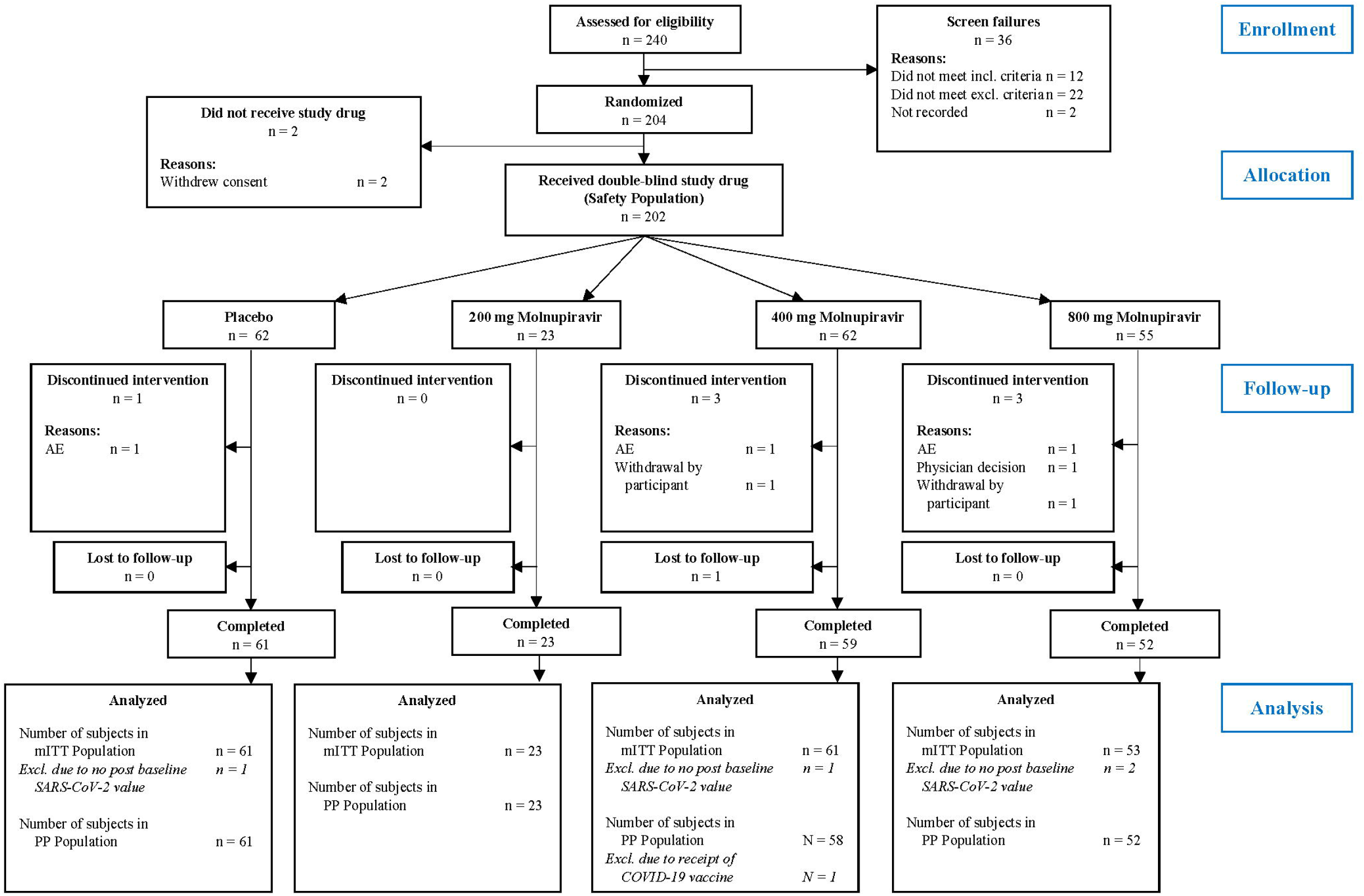
Study Flow Chart. Abbreviations: AE = adverse event; COVID-19 = coronavirus disease-2019; mITT = modified intent to treat; n = number of participants; PP = per protocol; SARS-CoV-2 = severe acute respiratory virus coronavirus 2. The mITT population included all subjects who were randomized into the study and had at least 1 post baseline viral RNA assessment. The PP population included subjects in the safety population who had no important protocol deviations leading to exclusion from the PP population and had completed the Day 28 follow-up visit. The safety population included all subjects who were randomized and took at least 1 dose of study drug. Subjects were analyzed according to the treatment they actually received.

### Isolation of Infectious SARS-CoV-2 Virus

Infectious virus was isolated from 43.5% (74/170) of evaluable nasopharyngeal swabs at baseline (Table 2 and Figure 2a). On Day 3, infectious virus isolation decreased significantly to 1.9% (1/53) of participants administered 800 mg molnupiravir compared to 16.7% (9/54) of participants administered placebo (p = 0.02). Infectious virus isolation also decreased on Day 5 in participants administered 400 or 800 mg molnupiravir, with zero having infectious virus isolated in these groups (0/42 and 0/53 respectively) compared to 11.1% (6/54) of placebo recipients (p = 0.03). The difference in infectious virus isolation remained significant for 400 and 800 mg molnupiravir groups (compared to placebo) on Day 5 when analysis was limited to those who were antibody negative at baseline and/or those with infectious virus isolation present at baseline (Supplementary Table 3 and Supplementary Table 4). The difference in infectious virus isolation remained significant for 800 mg molnupiravir when compared to concurrent placebo recipients (Supplementary Table 5). Overall, there was a clear dose response in infectious virus isolation, with the proportion of participants with infectious virus isolated significantly lower in the 400 or 800 mg molnupiravir compared to 200 mg molnupiravir or placebo on Days 3 and 5 (p-values = 0.010 and 0.003, respectively; Table 2) and the dose response was significant on Day 5 when analysis was limited to participants who had infectious virus isolation at baseline (p-value = 0.004; Supplementary Table 3).

**Table 2.** Summary of SARS-CoV-2 Infectivity and Virology.

| **Percentage of Participants Positive for Infectious SARS-CoV-2 Virus** |  |  |  |  |
| --- | --- | --- | --- | --- |
|  | **200 mg** | **400 mg** | **800 mg** | **Placebo** |
|  | **Molnupiravir** | **Molnupiravir** | **Molnupiravir** |  |
| Day 1, n/N (%) | 11/22 (50.0) | 18/43 (41.9) | 20/52 (38.5) | 25/53 (47.2) |
| Day 3, n/N (%) | 4/22 (18.2) | 5/43 (11.6) | 1/53 (1.9) | 9/54 (16.7) |
| *Fisher's exact p-value* | >0.99 | 0.57 | 0.016 |  |
| *Dose response p-value* |  |  |  | 0.010 |
| Day 5, n/N (%) | 1/22 (4.5) | 0/42 (0.0) | 0/53 (0.0) | 6/54 (11.1) |
| *Fisher's exact p-value* | 0.67 | 0.03 | 0.03 |  |
| *Dose response p-value* |  |  |  | 0.003 |
| **Time to SARS-CoV-2 Viral RNA Negativity** |  |  |  |  |
|  | **200 mg** | **400 mg** | **800 mg** | **Placebo** |
|  | **Molnupiravir** | **Molnupiravir** | **Molnupiravir** |  |
| Participants with Response, n/N (%) | 21/23 (91.3) | 48/61 (78.7) | 49/53 (92.5) | 49/61 (80.3) |
| Median time to response (95% CI), days | 22.0 (15.0, 28.0) | 27.0 (15.0, 28.0) | 14.0 (13.0, 14.0) | 15.0 (15.0, 27.0) |
| *Log-rank p-value* | 0.56 | 0.73 | 0.013 |  |

| Change from Baseline in SARS-CoV-2 Viral Load (log<sub>10</sub> copies/mL) |  |  |  |  |
| --- | --- | --- | --- | --- |
|  | 200 mg | 400 mg | 800 mg | Placebo |
|  | Molnupiravir | Molnupiravir | Molnupiravir |  |
| Day 3, n/N | 23/23 | 58/61 | 51/53 | 56/61 |
| *Least squares mean (SE)* | -0.783 (0.189) | -0.941 (0.118) | -1.050 (0.115) | -0.847 (0.127) |
| *Difference in least squares mean* | 0.064 | -0.094 | -0.203 |  |
| *95% CI* | -0.397, 0.525 | -0.438, 0.250 | -0.543, 0.137 |  |
| *p-value* | 0.78 | 0.59 | 0.24 |  |
| Day 5, n/N | 23/23 | 56/61 | 52/53 | 57/61 |
| *Least squares mean (SE)* | -1.471 (0.212) | -1.754 (0.128) | -1.867 (0.126) | -1.320 (0.150) |
| *Difference in least squares mean* | -0.150 | -0.434 | -0.547 |  |
| *95% CI* | -0.674, 0.374 | -0.825, -0.043 | -0.935, -0.159 |  |
| *p-value* | 0.57 | 0.030 | 0.006 |  |
| Day 7, n/N | 23/23 | 51/61 | 49/53 | 56/61 |
| *Least squares mean (SE)* | -2.028 (0.202) | -2.263 (0.119) | -2.485 (0.107) | -1.952 (0.157) |
| *Difference in least squares mean* | -0.076 | -0.311 | -0.534 |  |
| *95% CI* | -0.591, 0.438 | -0.702, 0.079 | -0.910, -0.157 |  |
| *p-value* | 0.77 | 0.12 | 0.006 |  |
| Day 14, n/N | 23/23 | 53/61 | 48/53 | 54/61 |
| *Least squares mean (SE)* | -2.884 (0.091) | -2.840 (0.065) | -3.040 (0.040) | -2.865 (0.106) |
| *Difference in least squares mean* | -0.019 | 0.026 | -0.175 |  |
| *95% CI* | -0.298, 0.260 | -0.220, 0.272 | -0.400, 0.050 |  |
| *p-value* | 0.89 | 0.84 | 0.13 |  |

Abbreviations: CI = confidence interval; n = number of observations; N = number of participants; SE = standard error.

**Figure 2.**
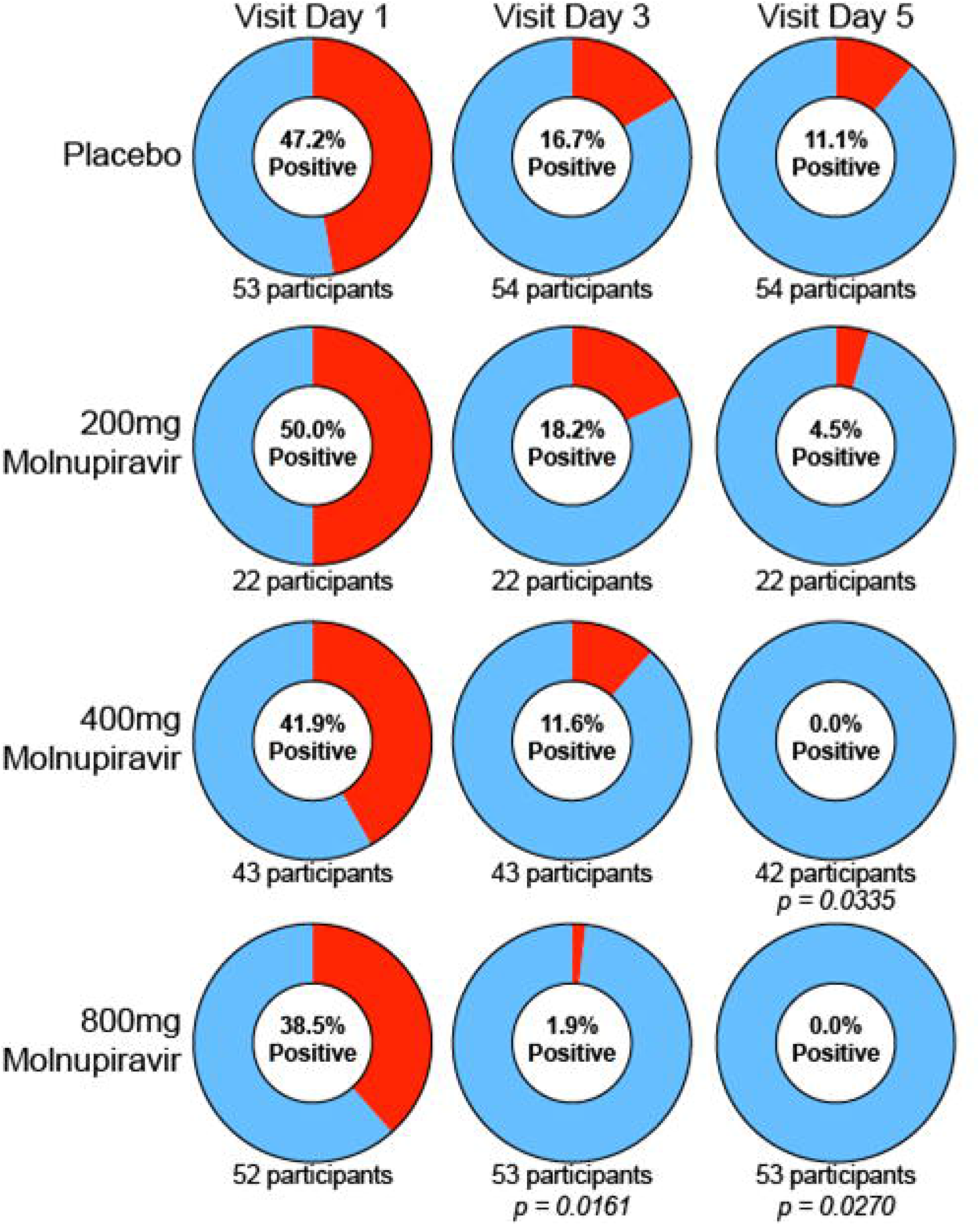
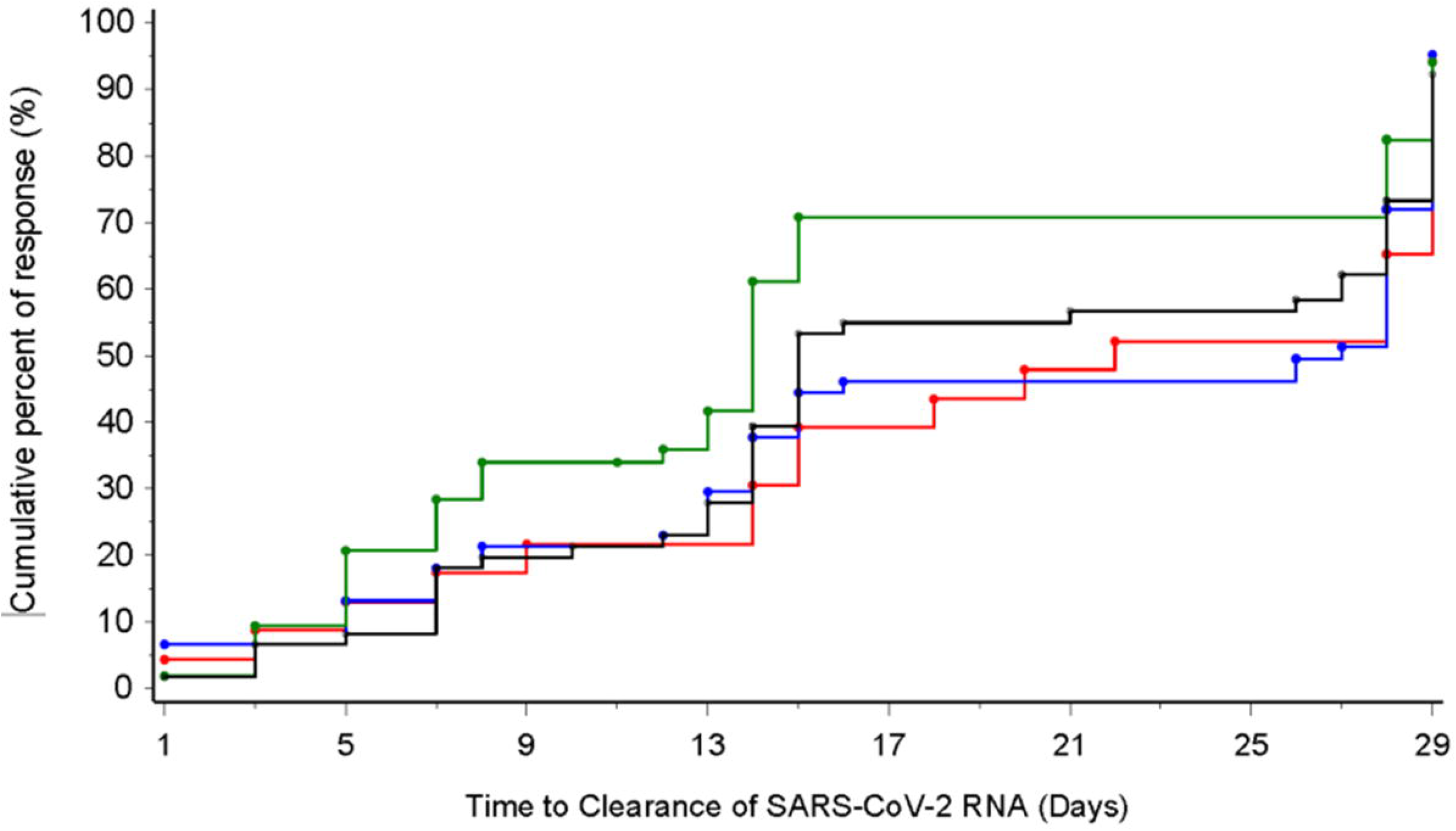
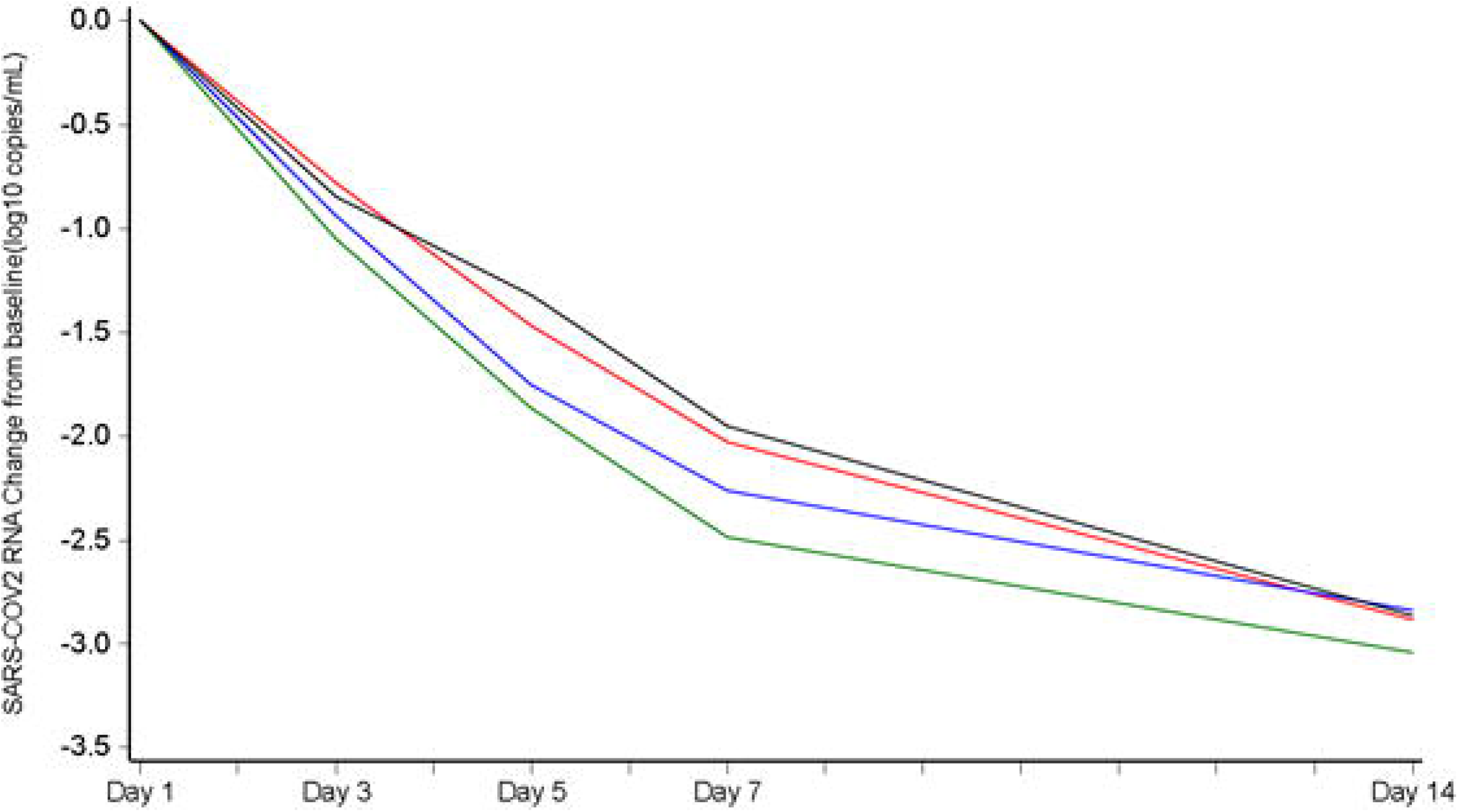
SARS-CoV-2 Infectivity and Virology. **(A)** Proportion of participants positive (red) for SARS-CoV-2 infectious virus; participants who are negative for SARS-CoV-2 infectious virus are in blue. **(B)** Kaplan-Meier plot of time to clearance of SARS-CoV-2 RNA by treatment (200 mg molnupiravir [red], 400 mg molnupiravir [blue], 800 mg molnupiravir [green], and placebo [black]). **(C)** Least squares mean change from baseline in SARS-CoV-2 RNA (log_10_ copies/mL) for the modified intent to treat population (200 mg molnupiravir [red], 400 mg molnupiravir [blue], 800 mg molnupiravir [green], and placebo [black]). Abbreviations: RNA = ribonucleic acid; SARS-CoV-2 = severe acute respiratory syndrome 2 coronavirus 2.

### Time to SARS-CoV-2 Clearance of Viral RNA

Time to clearance of viral RNA in nasopharyngeal swabs was the primary endpoint of this study and was significantly reduced in participants receiving 800 mg molnupiravir (median: 14 days) compared to those administered placebo (log-rank p-value = 0.013; Table 2 and Figure 2b). The reduction in time to clearance of viral RNA between 800 mg molnupiravir and placebo was greater and remained significant (medians: 14 and 27 days, respectively; p-value = 0.001) when analysis was limited to participants who were negative for antibodies at baseline (Supplementary Table 7). The proportion of participants who achieved SARS-CoV-2 negativity by end of study was also greater for those administered 800 mg molnupiravir (92.5%) compared with 91.3%, 78.7%, and 80.3% for 200 and 400 mg molnupiravir and placebo, respectively.

### Change from Baseline in SARS-CoV-2 Viral Load

The decrease in viral RNA from baseline to Days 3 to 28 was greater for the 800 mg molnupiravir group than any other group (Table 2 and Figure 2c). For participants administered 400 or 800 mg molnupiravir, the least squares mean viral load change from baseline was significantly greater on Day 5 when compared to placebo, with differences in least squares means of -0.434 and -0.547 log_10_ copies/mL (p = 0.03 and 0.006), respectively. Additionally, for participants administered 800 mg molnupiravir, the least squares mean viral load change from baseline was also significantly greater on Day 7 compared to placebo, with a least squares mean difference of -0.534 log_10_ copies/mL (p = 0.006). The reduction in viral load from baseline to Day 5 between 800 mg molnupiravir and placebo remained significant during sensitivity analyses of participants who were negative for antibodies at baseline (least squares mean difference of -0.613 log_10_ copies/mL; p = 0.002; Supplementary Table 6) and when compared to concurrent placebo (least squares mean difference of -0.376 log_10_ copies/mL; p = 0.045; Supplementary Table 8).

### SARS-CoV-2 Antibody Detection

Participants were tested for SARS-CoV-2-specific immunoglobulin (Ig) A, IgM, and IgG at baseline and on Days 7 and 28. The proportions of participants with any antibody to SARS-CoV-2 at baseline varied between the groups, with 15.0%, 30.0%, 35.3%, and 18.2% in the 200, 400, 800 mg molnupiravir and placebo groups respectively. By Day 28, 99.2% of molnupiravir-treated participants had developed antibodies to SARS-CoV-2, compared to 96.5% of those administered placebo.

### Next Generation Sequencing of SARS-CoV-2 RNA-Dependent RNA Polymerase

Genotypic changes in the RdRp occurred at a higher rate among participants who received molnupiravir compared to placebo. On average, 5.7 nucleotide changes in the RdRp were observed following treatment with placebo. By comparison, the average number of nucleotide changes in RdRp following molnupiravir treatment was two-fold greater (10.9) and was statistically significant (p = 0.02) supporting viral error catastrophe as the mechanism of action.^11^

### Safety and Tolerability

Molnupiravir and placebo were associated with few, and mainly low-grade, adverse events (Table 3). The incidence of treatment-associated adverse events was lowest in the molnupiravir 800 mg group. The only adverse events reported by more than 4 participants were headache, insomnia, and increased alanine aminotransferase. Two (1.4%) adverse events led to discontinuation of molnupiravir compared with 1 (1.6%) for placebo. Grade 3 or higher adverse events occurred in 5.0% and 8.1% in the molnupiravir and placebo groups, respectively. There were no treatment- or dose-related trends in hematology or clinical chemistry data during the study.

**Table 3.** Summary of Adverse Events.

| **Number (%) of participants experiencing**<br>**an event** | **Molnupiravir 200 mg**<br>**N = 23** | **Molnupiravir 400 mg**<br>**N = 62** | **Molnupiravir 800 mg**<br>**N = 55** | **Placebo**<br>**N = 62** |
| --- | --- | --- | --- | --- |
| Any adverse event | 11 (47.8) | 20 (32.3) | 11 (20.0) | 18 (29.0) |
| Adverse events reported by >5% subjects in any group |  |  |  |  |
| Dizziness | 2 (8.7) | 1 (1.6) | 0 | 0 |
| Insomnia | 2 (8.7) | 1 (1.6) | 1 (1.8) | 4 (6.5) |
| Any adverse event grade 3 or higher | 1 (4.3) | 2 (3.2) | 4 (7.3) | 5 (8.1) |
| Any adverse event leading to discontinuation<br>from study drug | 0 | 1 (1.6) | 1 (1.8) | 1 (1.6) |
| Any serious adverse event | 0 | 2 (3.2) | 1 (1.8) | 1 (1.6) |
| Any adverse event leading to death | 0 | 0 | 0 | 1 (1.6)* |

Abbreviation: n = number of participants.
\* The subject had an adverse event of hypoxia that led to death. This occurred 31 days after discontinuation from the study following completion of study assessments and was not recorded in the study database but was recorded in the safety database.

Four serious adverse events occurred and resulted in hospitalization, comprising one (1.6%) participant administered placebo who had hypoxia, two (3.2%) participants administered 400 mg molnupiravir (cerebrovascular accident and decreased oxygen saturation), and one (1.8%) participant administered 800 mg molnupiravir who had acute respiratory failure (Table 3). Treatment was discontinued in all 4 participants. Following completion, 1 death due to COVID-19 occurred in a participant administered placebo outside of the 28-day time window.

### Clinical Endpoints

A greater proportion of participants administered molnupiravir reported their health as poor or fair (68.2%) at baseline than those administered placebo (50.0%). There were no notable differences in symptom duration or severity between the treatment groups over the 28 days.

## Discussion

In this Phase 2a, randomized, double-blind, placebo-controlled, clinical trial, molnupiravir was well tolerated and associated with potent antiviral efficacy as evidenced by significantly reduced infectious virus isolation, time to elimination of SARS-CoV-2 RNA, increased proportion of participants that cleared SARS-CoV-2 RNA, and a greater reduction in SARS-CoV-2 viral RNA from baseline compared to placebo in outpatients with COVID-19. Four days after treatment initiation, there was no infectious virus isolated from any participants who received 400 or 800 mg molnupiravir.

The reduction in the time to clearance of viral RNA, the greater proportion of participants with SARS-CoV-2 viral RNA negativity, and the greater change in viral RNA levels from baseline in participants treated with 800 mg molnupiravir compared to placebo support the findings of a significant decrease in infectious virus isolation. These results remained significant even when accounting for time from symptom onset, viral RNA level, and seropositivity. Importantly, only 22.8% of participants had evidence of a humoral immune response at baseline, indicating that randomization occurred early in the clinical course of infection. However, by Day 28, 92.1% were antibody positive, demonstrating that early treatment with molnupiravir had an antiviral effect without inhibiting the development of a humoral immune response.

In clinical trials of COVID-19 therapeutics, antiviral efficacy against SARS-CoV-2 has been assessed by measuring viral RNA levels; however, detection of viral RNA does not confirm the replication competence of the virus. We believe demonstration of the ability of treatment to reduce and eliminate infectious SARS-CoV-2 is an essential consideration; in observational studies, SARS-CoV-2 virus isolation decreases with time, and persistence of infectious virus is associated with disease severity and host immune status.^4,20,21^ In otherwise healthy adults with mild to moderate disease, infectious virus has been isolated up to 10 days after symptom onset.^3,7,20,21^ In this, the largest outpatient study to evaluate infectious virus isolation to date, infectious SARS-CoV-2 was isolated from 11.1% of placebo recipients at study Day 5, a mean 9 to 10 days from symptom onset, and treatment with molnupiravir was found to accelerate the clearance of infectious virus. Critically, this is the first clinical trial to demonstrate the ability of treatment to reduce and eliminate infectious SARS-CoV-2 in patients with COVID-19.

Safety analyses from this trial were consistent with those from a Phase 1 trial of molnupiravir and support ongoing clinical development.^9^ Overall, molnupiravir was well tolerated with no increase in treatment-related or serious adverse events compared to participants administered placebo. There were no safety signals or evidence of hematologic, renal, or hepatic toxicity at any dose.

Importantly, this clinical trial was designed to evaluate the antiviral efficacy of molnupiravir and was not powered to evaluate clinical endpoints, such as symptom duration or hospitalization. However, a Phase 2/3 study is ongoing to evaluate the effect of molnupiravir on symptom duration and severity, emergency department visits, and hospitalizations (ClinicalTrials.gov Identifier: NCT04575597). Another limitation included imbalances in the randomization, with a greater proportion of seropositive individuals and a trend toward a lower viral load at baseline among those randomized to 800 mg molnupiravir compared to placebo. However, differences in infectious virus isolation at Day 5, time to clearance of viral RNA, and reductions in viral load from baseline to Day 5 remained significant when analysis was limited to participants who were antibody negative at baseline. Among participants who were negative for antibodies at baseline, there was no difference in baseline viral load between the 800 mg molnupiravir and placebo groups. An additional limitation is the use of pooled placebo in the analyses; however, differences in virus isolation and viral load reduction on Day 5 remained significant when analysis was limited to comparisons between 800 mg molnupiravir and concurrent placebo (Supplementary Table 7). These sensitivity analyses indicate that the antiviral efficacy demonstrated in this study is not due to imbalances in either seropositivity, trends in viral RNA level at baseline, or the use of a pooled placebo group.

Currently, two combination intravenous monoclonal antibody cocktails have been granted emergency use authorization in an attempt to reduce nasopharyngeal SARS-CoV-2 RNA, medical visits, and hospitalizations; however, the logistical challenges of intravenous therapies limit timely administration to patients who may benefit most from these important treatments.^2,22–24^ This trial provides strong biological evidence that supports development of molnupiravir as an oral agent to reduce infectious virus replication and interrupt progression of COVID-19 in early stages of disease. Current evidence suggests that uninterrupted viral replication is a major sign of progression to more severe disease.^2,5^ Critically, molnupiravir can be produced at scale and does not require cold transportation or infection control infrastructure for administration.

In conclusion, the results of this trial demonstrate the safety, tolerability, and antiviral efficacy of molnupiravir to reduce replication of SARS-CoV-2 and accelerate clearance of infectious virus and support ongoing trials of molnupiravir to prevent progression of COVID-19 and eliminate onward transmission of SARS-CoV-2.^8^

## Data Availability

The data referred to in this manuscript will be available within 1 year of study completion.

## Acknowledgements

The authors with to acknowledge the participants and the following sites for their dedication and contributions to the study: Valley Clinical Trials (Northridge, CA), University of North Carolina (Chapel Hill, NC), Fred Hutchinson (Seattle, WA), Care United Research (Forney, TX), Benchmark Research (Colton, CA), FOMAT Medical Research (Oxnard, CA), Indago Research & Health Center (Hiahleah, FL), Wake Forest University (Winston-Salem, NC), Duke University (Durham, NC), and NOLA Research Works (New Orleans, LA). Additionally, the authors with to acknowledge Dr Mark Stead of Covance for supporting the preparation of this manuscript and the randomization support provided by the National Center for Advancing Translational Sciences (NCATS; UL1TR002489) and the UNC Center for AIDS Research (P30 AI050410). Molnupiravir was invented at Drug Innovations at Emory (DRIVE) LLC, a not-for-profit biotechnology company wholly owned by Emory University, and with partial funding support from the US government. Since licensed by Ridgeback Biotherapeutics, all funds used for the development of molnupiravir by Ridgeback Biotherapeutics have been provided by Wayne and Wendy Holman and Merck.

## Notes

### Competing Interest Statement

All authors have completed the ICMJE uniform disclosure form at www.icmje.org/coi_disclosure.pdf and declare the following: PA, FL, WP, LS, and WH were employed by Ridgeback Biotherapeutics in direct support of the work reported in this manuscript. MA, WF, KM, and RB were contracted by Ridgeback Biotherapeutics in direct support of the work reported in this manuscript. JE, WP, TS, DW, CW, and WF are independent contractors who were topically related to this study within the past 36 months. MC was employed by UNC at Chapel Hill and received a grant from the National Institute of Health in direct support of the work presented in this manuscript. KM has stocks or stock options in ICON Plc. WP and WH received royalties from patents, trademarks, copyrights or other intellectual property. WH has stock or stock options and is a fiduciary offer or member of another board. LF, KB-E, AJL and ED declared no conflicts of interest. The initial draft of this manuscript was prepared by Mark Stead of Covance Clinical Research Unit Limited, UK.

### Clinical Trial

NCT04405570

### Author Declarations

The protocol was approved by: WCG IRB 1019 39th Ave SE, Suite 120 Puyallup, WA 98374

