## Supplementary Information for "Molnupiravir, an Oral Antiviral Treatment for COVID-19"

**Supplementary Methods**

Sample Size Determination	2

Blinding	3

Randomization	3

Handling of Missing Data and Outliers	4

Analysis Methods	4

**Supplementary Tables**

Supplementary Table 1. Missing Data for SARS‑CoV‑2 Infectious Virus	5

Supplementary Table 2. Missing Data for Time to SARS‑CoV‑2 Viral RNA Negativity	6

Supplementary Table 3. Summary of SARS‑CoV‑2 Infectivity for Participants Who Had Infectious Virus Isolation at Baseline	7

Supplementary Table 4. Summary of SARS‑CoV‑2 Infectivity for Participants Who Had Infective Virus Isolation at Baseline and Were Negative for Antibodies	8

Supplementary Table 5. Summary of SARS‑CoV‑2 Infectivity for Participants Who Were Administered 800 mg Molnupiravir Versus Concurrent Placebo	9

Supplementary Table 6. Change from Baseline in SARS‑CoV‑2 Viral Load (log_10_ copies/mL) in Participants Who Were Negative for Antibodies at Baseline	10

Supplementary Table 7. Time to SARS‑CoV‑2 Viral RNA Negativity in Participants Who Were Negative for Antibodies at Baseline	12

Supplementary Table 8. Change from Baseline in SARS‑CoV‑2 Viral Load (log_10_ copies/mL)	13

Sample Size Determination

The primary endpoint was evaluated using a Kaplan‑Meier estimation with a corresponding exact log‑rank test. Anticipating up to 10% missing data, 44 participants (approximately 22 for each active and placebo) provided 88% power to detect a risk difference (active – placebo) of ‑35% in the primary endpoint using a type I error rate (alpha) of 0.10. The sample size in the dose escalation part was based on assessment of the primary endpoint across all parts. A total of 96 active and 32 placebo would achieve 83% power to detect a between‑group difference of ≥0.6 standard deviations, with a type I error rate (alpha) of 0.05 using a two‑sided test.

Blinding

The investigator, subjects, and sponsor were blinded to the treatments received. However, a subset of sponsor representatives was unblinded to the treatment randomization to facilitate ongoing review of virology data.

Randomization

Participants were randomized using the REDCap randomization application (The University of North Carolina and Chapel Hill Translational and Clinical Sciences Institute and REDCap Development Team, The University of North Carolina and Chapel Hill, NC).

The randomization schedule was generated using PROC PLAN in SAS Version 9.4 (SAS Institute Inc., Cary NC) by an independent statistician. Randomization was at a 1:1 ratio (molnupiravir:placebo) in Part 1 and was stratified by time since onset of coronavirus disease‑2019 (COVID‑19) symptoms (early [0 to ≤60 hours] versus late [>60 to ≤168 hours]). Randomization was at a 3:1 ratio (molnupiravir:placebo) in Parts 2 to 9 and was not stratified.

The randomization in Part 1 used 6 blocks of size 2 plus 8 blocks of size 4 to generate a total of 44 allocations to each of the early and late strata, in case either of the strata enrolled all of the participants in Part 1. An additional 44 randomization allocations were appended to the end of each stratum (for a total of 88 in each stratum) using the same permuted block design to allow for approved replacement or over‑enrollment. The randomization in Parts 2 to 9 used a fixed block size of 4 to generate either 4 blocks of size 4, 5 blocks of size 4, or 6 blocks of size 4 (ie, 16, 20, or 24 participants), depending on the requested number of assignments. An additional 4 randomization allocations were appended to each part (for a total of 20 across Parts 2 to 9) for approved replacement or over‑enrollment. Within‑ and between‑block seeds were randomly generated in SAS Version 9.4 prior to initialization of the random number generator.

Handling of Missing Data and Outliers

Participants with missing infectivity and SARS‑CoV‑2 RNA data were imputed based on the algorithms provided in Supplementary Table 1 and Supplementary Table 2.

Analysis Methods

Time to viral RNA clearance was defined as the first time viral RNA achieved and maintained below the lower limit of quantitation (<1,018 copies/mL). If the first negative test occurred on the last on study assessment, it was considered achieving the viral RNA clearance on the last assessment. Participants who completed the study without achieving viral RNA clearance were censored at the last viral RNA assessment. Participants who discontinued the study without achieving viral RNA clearance were censored at Day 28. Three nasopharyngeal swabs (2 from 400 mg molnupiravir‑treated participants who tested negative and 1 from a placebo participant who tested positive) were analyzed despite having been received out of temperature range.

Additional information on the conduct of the trial and analyses of data are presented in the protocol, protocol amendments, and statistical analysis plans that have been made available alongside this manuscript.

Supplementary Table 1. Missing Data for SARS‑CoV‑2 Infectious Virus

| **Visit Before** | **Visit After** | **Imputation of Infectivity Result** |
| --- | --- | --- |
| Negative | Negative | Negative |
| Positive | Positive | Positive |
| Negative or missing | Positive | Positive* |
| Missing | Negative | Missing |
| Any value | Missing | Missing |

* For participants with missing baseline assessments and reported the first post baseline value as positive, the baseline was imputed as positive.

Supplementary Table 2. Missing Data for Time to SARS‑CoV‑2 Viral RNA Negativity

| **Visit Before** | **Visit After** | **Imputation of Undetectable SARS‑CoV‑2 RNA** |
| --- | --- | --- |
| BLQ/BLD | BLD or BLQ | Negative |
| >BLQ | BLD or BLQ | Positive |
| BLD/BLQ or missing | >BLQ | Positive* |
| Missing | BLD or BLQ | Missing |
| Any value | Missing | Missing |

Abbreviations: BLD = below limit of detection; BLQ = below limit of quantification.

* For participants with missing baseline assessments and reported the first post baseline value as >BLQ, the baseline was imputed as >BLQ (positive).

Supplementary Table 3. Summary of SARS‑CoV‑2 Infectivity for Participants Who Had Infectious Virus Isolation at Baseline

|  | **200 mg Molnupiravir**  N = 11 | **400 mg Molnupiravir**  N = 18 | **800 mg Molnupiravir**  N = 20 | **Placebo**  N = 25 |
| --- | --- | --- | --- | --- |
| Day 3, n/N (%) | 4/11 (36.4) | 5/18 (27.8) | 1/20 (5.0) | 7/25 (28.0) |
| *Fisher’s exact p‑value* | 0.70 | >0.99 | 0.06 |  |
| *Dose response p‑value* |  |  |  | 0.06 |
| Day 5, n/N (%) | 1/11 (9.1) | 0/18 (0.0) | 0/20 (0.0) | 6/25 (24.0) |
| *Fisher’s exact p‑value* | 0.40 | 0.03 | 0.03 |  |
| *Dose response p‑value* |  |  |  | 0.004 |

Abbreviations: n = number of observations; N = number of participants.

Supplementary Table 4. Summary of SARS‑CoV‑2 Infectivity for Participants Who Had Infective Virus Isolation at Baseline and Were Negative for Antibodies

|  | **200 mg Molnupiravir**  **N = 9** | **400 mg Molnupiravir**  **N = 17** | **800 mg Molnupiravir N = 20** | **Placebo**  **N = 24** |
| --- | --- | --- | --- | --- |
| Day 3, n/N (%) | 2/9 (22.2) | 5/17 (29.4) | 1/20 (5.0) | 7/24 (29.2) |
| *Fisher’s exact p‑value* | >0.99 | >0.99 | 0.05 |  |
| *Dose response p‑value* |  |  |  | 0.07 |
| Day 5, n/N (%) | 0/9 (0.0) | 0/17 (0.0) | 0/20 (0.0) | 6/24 (25.0) |
| *Fisher’s exact p‑value* | 0.16 | 0.03 | 0.02 |  |
| *Dose response p‑value* |  |  |  | 0.003 |

Abbreviations: n = number of observations; N = number of participants.

Supplementary Table 5. Summary of SARS‑CoV‑2 Infectivity for Participants Who Were Administered 800 mg Molnupiravir Versus Concurrent Placebo

| Number of Positive Subjects | **800 mg Molnupiravir** | **Placebo** |
| --- | --- | --- |
| Day 1, n/N (%) | 20/52 (38.5%) | 8/17 (47.1) |
| *p‑value* | 0.58 |  |
| Day 3, n/N (%) | 1/53 (1.9) | 2/18 (11.1) |
| *p‑value* | 0.16 |  |
| Day 5, n/N (%) | 0/53 (0.0) | 3/18 (16.7) |
| *p‑value* | 0.01 |  |
| Day 7, n/N (%) | 0/52 (0.0) | 1/18 (5.6) |
| *p‑value* | 0.26 |  |

Abbreviations: n = number of observations; N = number of participants.

Supplementary Table 6. Change from Baseline in SARS‑CoV‑2 Viral Load (log_10_ copies/mL) in Participants Who Were Negative for Antibodies at Baseline

|  | **200 mg Molnupiravir** | **400 mg Molnupiravir** | **800 mg Molnupiravir** | **Placebo** |
| --- | --- | --- | --- | --- |
| Day 3, n/N | 17/17 | 34/34 | 31/32 | 44/44 |
| *Least squares mean (SE)* | ‑1.141 (0.183) | ‑1.196 (0.132) | ‑1.157 (0.129) | ‑0.877 (0.150) |
| *Difference in least squares mean* | ‑0.263 | ‑0.319 | ‑0.280 |  |
| *95% CI* | ‑0.734, 0.207 | ‑0.714, 0.076 | ‑0.670, 0.111 |  |
| *p‑value* | 0.27 | 0.11 | 0.16 |  |
| Day 5, n/N | 17/17 | 34/34 | 31/32 | 44/44 |
| *Least squares mean (SE)* | ‑1.874 (0.183) | ‑2.101 (0.132) | ‑2.191 (0.129) | ‑1.578 (0.150) |
| *Difference in least squares mean* | ‑0.296 | ‑0.523 | ‑0.613 |  |
| *95% CI* | ‑0.767, 0.174 | ‑0.917, ‑0.128 | ‑1.004, ‑0.223 |  |
| *p‑value* | 0.22 | 0.010 | 0.002 |  |
| Day 7, n/N | 17/17 | 31/34 | 31/32 | 43/44 |
| *Least squares mean (SE)* | ‑2.447 (0.183) | ‑2.582 (0.137) | ‑2.967 (0.129) | ‑2.284 (0.152) |
| *Difference in least squares mean* | ‑0.163 | ‑0.298 | ‑0.683 |  |
| *95% CI* | ‑0.635, 0.308 | ‑0.702, 0.105 | ‑1.075, ‑0.290 |  |
| *p‑value* | 0.50 | 0.15 | <0.001 |  |
| Day 14, n/N | 17/17 | 32/34 | 31/32 | 41/44 |
| *Least squares mean (SE)* | ‑3.579 (0.183) | ‑3.412 (0.136) | ‑3.601 (0.129) | ‑3.343 (0.155) |
| *Difference in least squares mean* | ‑0.236 | ‑0.069 | ‑0.257 |  |
| *95% CI* | ‑0.711, 0.239 | ‑0.475, 0.337 | ‑0.654, 0.140 |  |
| *p‑value* | 0.33 | 0.74 | 0.20 |  |

Abbreviations: CI = confidence interval; n = number of observations; N = number of participants; SE = standard error.

Supplementary Table 7. Time to SARS‑CoV‑2 Viral RNA Negativity in Participants Who Were Negative for Antibodies at Baseline

|  | **200 mg Molnupiravir** | **400 mg Molnupiravir** | **800 mg Molnupiravir** | **Placebo** |
| --- | --- | --- | --- | --- |
| Participants with Response, n/N (%) | 16/17 (94.1) | 26/34 (76.5) | 32/32 (100) | 34/44 (77.3) |
| Median time to response (95% CI), days | 22.0 (15.0, 29.0) | 26.0 (15.0, 28.0) | 14.0 (13.0, 14.0) | 27.0 (15.0, 28.0) |
| *Log‑rank p‑value* | 0.87 | 0.48 | 0.001 |  |

Abbreviations: CI = confidence interval; n = number of observations; N = number of participants.

Supplementary Table 8. Change from Baseline in SARS‑CoV‑2 Viral Load (log_10_ copies/mL)

|  | **800 mg Molnupiravir**  N = 53 | **Placebo**  N = 18 |
| --- | --- | --- |
| Day 3, n/N | 51/53 | 16/18 |
| *Least squares mean (SE)* | -0.955 (0.094) | -0.638 (0.166) |
| *Difference in least squares mean* | -0.316 |  |
| *95% CI* | -0.696, 0.063 |  |
| *p‑value* | 0.1010 |  |
| Day 5, n/N | 52/53 | 17/18 |
| *Least squares mean (SE)* | -1.673 (0.093) | -1.297 (0.160) |
| *Difference in least squares mean* | -0.376 |  |
| *95% CI* | -0.744, -0.008 |  |
| *p‑value* | 0.0453 |  |

Abbreviations: CI = confidence interval; n = number of observations; N = number of participants; SE = standard error.
